# DEEP LEARNING TISSUE ANALYSIS DIAGNOSES AND PREDICTS TREATMENT RESPONSE IN EOSINOPHILIC ESOPHAGITIS

**DOI:** 10.1101/2021.06.10.21258624

**Authors:** Aamir Javaid, Philip Fernandes, William Adorno, Alexis Catalano, Lubaina Ehsan, Hans Vizthum von Eckstaedt, Barrett Barnes, Marium Khan, Shyam S. Raghavan, Emily McGowan, Margaret H. Collins, Marc E. Rothenberg, Christopher A Moskaluk, Donald E. Brown, Sana Syed

**Affiliations:** University of Virginia School of Medicine, Charlottesville, VA, USA; University of Virginia School of Engineering, Charlottesville, VA, USA; University of Virginia School of Data Science, Charlottesville, VA, USA; Divisions of Pathology and Laboratory Medicine, University of Cincinnati, Cincinnati, Ohio, USA; Division of Allergy and Immunology, University of Cincinnati, Cincinnati, Ohio, USA

**Keywords:** EoE, deep learning, eosinophils, tissue features, treatment response

## Abstract

**Background:** Eosinophilic Esophagitis (EoE) is a chronic inflammatory condition diagnosed by ≥15 eosinophils (Eos) per high-power field (HPF). There is no gold standard for clinical remission and Eo-associated metrics are poorly correlated with symptoms. Deep learning can be used to explore the relationships of tissue features with clinical response.

**Objectives:** To determine if deep learning can elucidate tissue patterns in EoE that predict treatments or symptoms at remission.

**Methods:** We created two deep learning models using esophageal biopsies from histologically normal and EoE patients: one to identify Eos in esophageal biopsies and a second to broadly classify esophageal tissue as EoE vs. normal. We used these models to analyze biopsies at diagnosis and first remission timepoint, as defined by <15 Eos/HPF, in a subset of 19 treatment-naïve patients. Differences in deep learning metrics between patient groups were assessed using Wilcoxon Rank-Sum tests.

**Results:** All initial patients were symptomatic at diagnosis and a majority were still suffering from dysphagia at remission. The Eo identification model had a low mean (SD) error of −0.3 (11.5) Eos/HPF. Higher peak and average Eo counts at diagnosis were associated with higher likelihood of being on a food-elimination diet at remission than steroids or proton-pump inhibitor (p<0.05). The EoE classification model had an F1-score of 0.97 for distinguishing normal tissue from EoE. There was a significant decrease from diagnosis in the percentage of EoE-classified tissue among asymptomatic remission patients (p<0.05).

**Conclusions:** Deep learning may have utility in diagnosing EoE and predicting future treatment response at diagnosis and resolution of symptoms at follow-up.

**Clinical Implications or Key Messages (for mechanistic article):** We developed two deep learning approaches for tissue analysis in eosinophilic esophagitis, which may improve histologic assessment of patients at diagnosis and predict treatment response and symptoms at remission.

**Capsule summary:** Two deep learning approaches for eosinophilic esophagitis (EoE): (1) Quantification of eosinophils throughout an entire biopsy, which predicted treatment at remission (2) Classifying esophageal tissue as EoE or normal, which predicted symptoms at remission.

## INTRODUCTION

Eosinophilic esophagitis (EoE) is a relapsing chronic disorder diagnosed by symptoms of esophageal dysfunction and at least one high-powered field (HPF) with ≥15 eosinophils (Eos) on esophageal biopsy in the absence of other causes of esophageal eosinophilia.^1–6^ Although multiple consensus guidelines agree on standards for diagnosis, clinical trials have varied in the histologic, endoscopic, and clinical criteria for defining remission.^7,8^ Goals of therapy are often to reduce eosinophils <15 Eos/HPF and improve patient symptoms,^8^ but many patients who achieve <15Eos/HPF are still symptomatic.^9,10^ This intensifies a need for alternate metrics of remission, such as more stringent Eo-based criteria or histologic scoring systems;^8,11^ however, the bottleneck of human review may hinder the widespread adoption or efficacy of more comprehensive indices.

Deep learning can automate histological evaluation of EoE patients with the potential to reveal subtle interactions between tissue features that are related to patient-centered outcomes such as treatment response and resolution of symptoms.^12^ While recent publications have investigated use of deep learning for EoE patient evaluations,^13,14^ use of computer vision on histology to predict treatment or symptoms at remission remains unstudied. We developed two deep learning models – one that automatically identifies Eos throughout a patient’s entire biopsy whole-slide image (WSI) and a second that learns to broadly classify esophageal tissue as EoE versus normal – to determine if patterns in tissue morphology can predict clinically relevant outcomes and improve upon current EoE patient evaluation guidelines.

## METHODS

### Selection of participants

This retrospective study was conducted with approval from the University of Virginia (UVA) Institutional Review Board for Health Sciences Research (IRB-HSR; IRB #20448). The UVA Clinical Data Repository provided data for patients who had undergone esophagogastroduodenoscopy (EGD) and had histologically normal esophagus or EoE reported on their pathology reports. This study included EoE patients who were diagnosed in accordance with consensus guidelines – esophageal symptoms including dysphagia and food impaction with eosinophilic predominant inflammation of ≥15 eosinophils (Eos) in at least one high-powered field (HPF; 400x magnification adjustment).^1–6^ A total of 46 EoE patients were identified, of which 19 had initial diagnostic biopsy performed at UVA. A comparison group was selected of 41 patients with no disease in any part of the gastrointestinal tract. All EoE patients were not used for model training due to computational demands and limited human annotator time, as shown in Figure 1. Enough patches were used for training of both models to achieve high performance.

**Figure 1.**
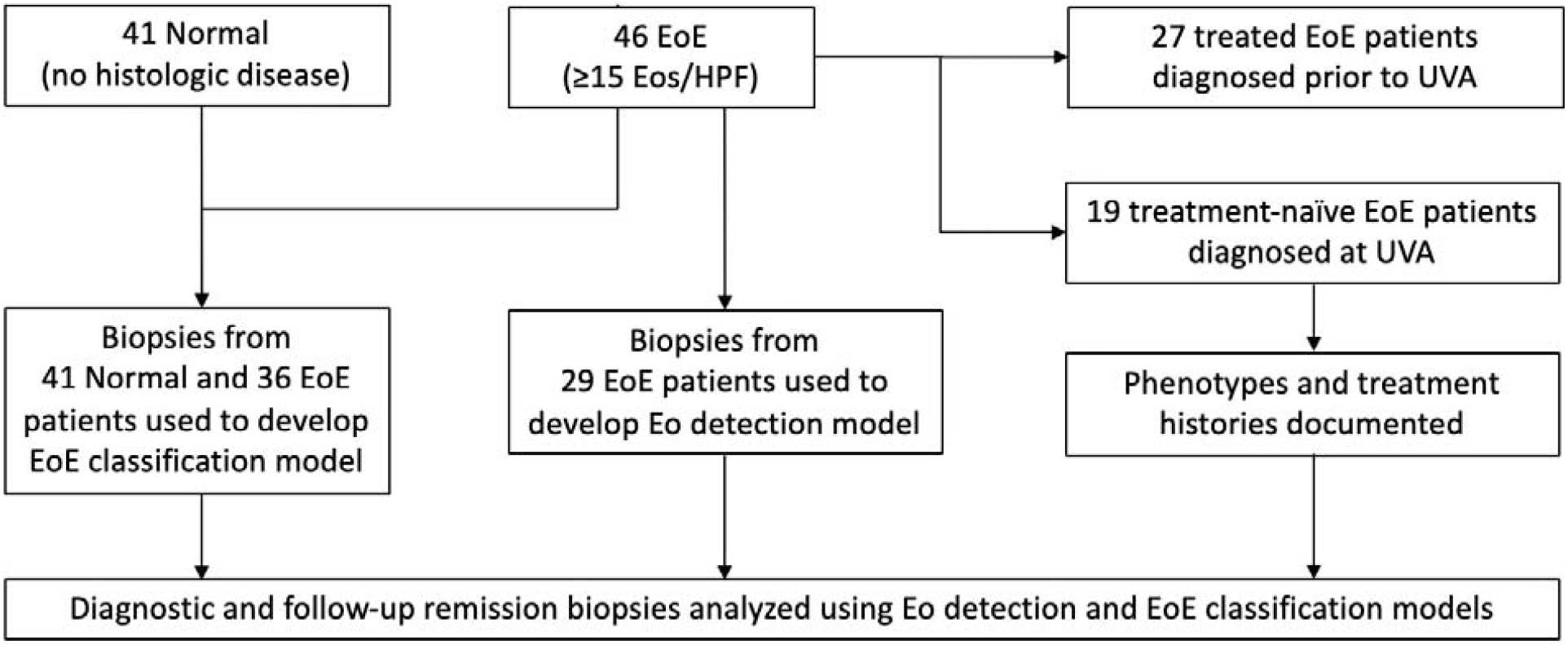
Selection of Participants. 41 histologically normal and 46 EoE patients with esophageal biopsies performed at UVA were located. 512×512 biopsy patches from all normal and a random-selection of 36 EoE patients (mix of those diagnosed at and before presenting to UVA) were used to develop the EoE classification model. Annotated biopsy patches from a random-selection of 29 EoE patients (mix of those diagnosed at and before presenting to UVA) were used to develop the Eo detection model. Of the 46 EoE patients, diagnostic and remission biopsies from 19 initially treatment-naïve patients were analyzed using both models. EoE=Eosinophilic esophagitis, UVA = University of Virginia.

### Creation of Deep Learning Models

To assist with the clinical assessment of EoE patients, we created two independent deep learning models. The first was an image segmentation model for automated detection of Eos across an entire esophageal histology slide. The second was a classification model which categorized esophageal tissue as EoE vs. histologically normal based on distinct tissue patterns identified by the model. Model development was an interdisciplinary collaboration between UVA’s medicine and engineering departments utilizing previously published pre- and post-processing steps.^15^

#### Developing Automated Eosinophil Detection Model

Esophageal biopsy slides from the study population were sent to the Biorepository and Tissue Research Facility (BTRF) at UVA where the Hematoxylin and Eosin (H&E) slides were digitized as whole slide images at approximately 100,000 by 30,000 pixels using a Hamamatsu NanoZoomer S360 Digital slide scanner C13220 (NanoZoomer S360 Digital slide scanner C13220-01, n.d.). The WSIs were pre-processed and cropped to 512 by 512-pixel patches for deep learning analysis. Whereas a standard 400x HPF usually measures around 0.21 mm^2^ or 2,000 by 2,000 pixels,^16^ smaller patches allowed for more detailed image annotation and reduced computational demands in training the models.^15^

Aperio ImageScope (Aperio ImageScope—Pathology Slide Viewing Software, n.d.) was used to annotate Eos in the patches used to train the Eo detection model. Annotations were performed by post-doctorate and medical trainees and reviewed by two board-certified gastrointestinal pathologists to ensure standardization between annotators. **Figure 2** shows an example of how Eos were annotated and converted to a ground truth mask for model training. A total of 513 annotated image patches from 29 EoE patients were used for model training (almost double the number of patches used in the previously published model).^15^

**Figure 2.**
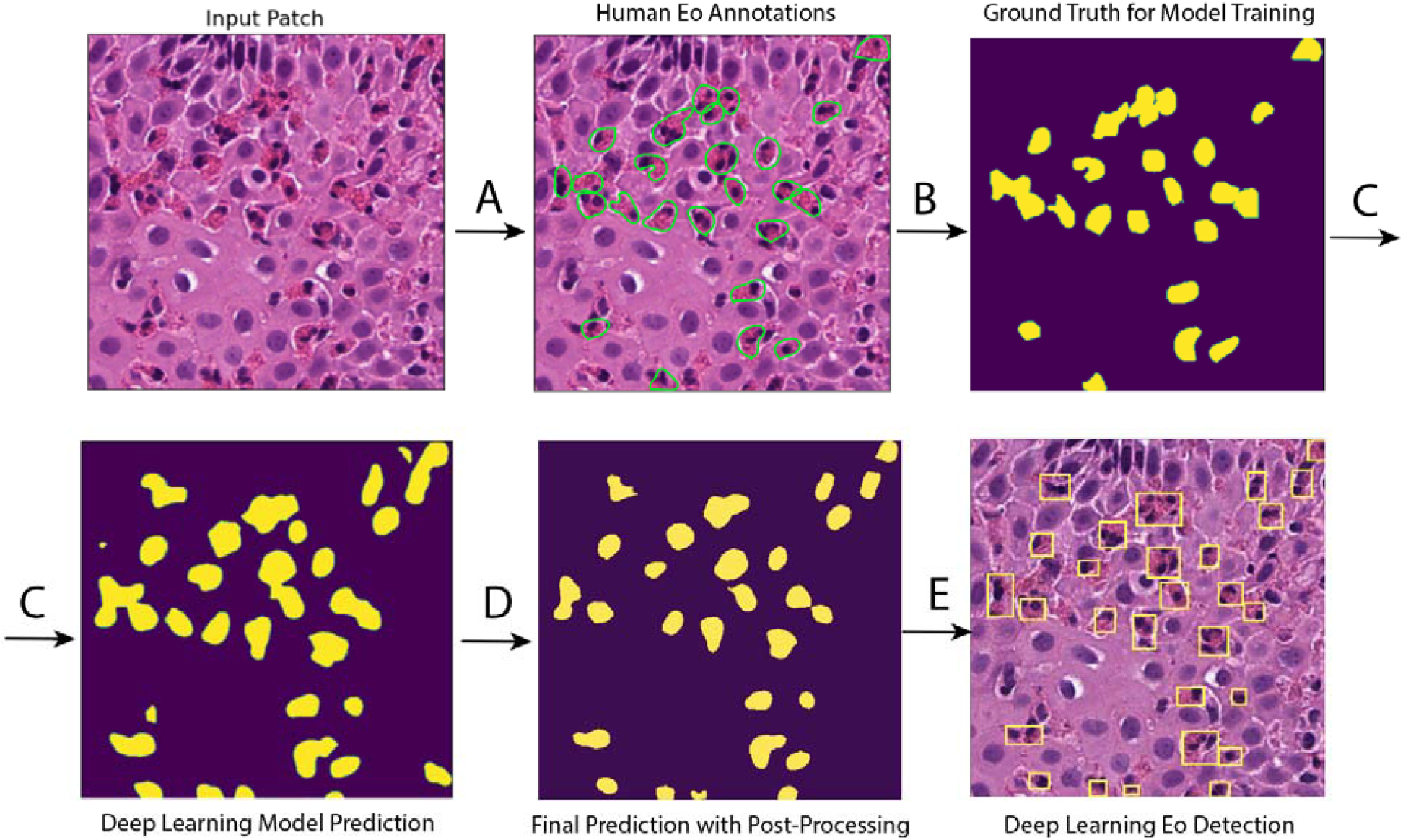
Eosinophil (Eo) detection model training. (A) Eos in 512×512 pixel biopsy patches were annotated by medical trainees with review by board-certified gastrointestinal pathologists. (B) 513 annotated patches were used to create ground truth masks for deep learning model training. (C) The model identifies features to distinguish parts of the image associated with Eos (yellow) from non-Eos (blue) and produces a prediction mask for all patches. (D) Prediction masks are post-processed to reduce overlap between adjacent cells. (E) Final deep learning Eo detection on original patch.

A U-Net model was developed to identify Eos in esophageal biopsies, as previously described.^15^ Deep learning prediction masks were post-processed, including use of a Euclidean distance transformation that reduced 8 pixels from the border of all Eos to minimize overlap between adjacent cells.^17^ We quantified the prediction accuracy using a 10-fold cross validation, where every image is rotated into the test set once, to assess the average counting error per patch, the error standard deviation, the Pearson correlation between true and predicted Eos, and the Dice coefficient.

#### Developing the EoE Classification Model

A second model was trained to classify biopsy images as EoE vs. histologically normal. 512 by 512-pixel patches from esophageal biopsies of histologically normal (n=41, 22,792 patches) and EoE patients (n=36, 26,949 patches) were used to develop a VGG16 Convolutional Neural Network (CNN) model to predict EoE versus normal tissue at a patch-level.^15^ Enough EoE patients were selected such that there were similar number of total patches for both the EoE and the histologically normal comparison group while each individual subject also contributed an equal number of patches to the model. Heterogeneity between histologically normal and EoE patients was assessed using a two-sample t-test for age and chi-square test for sex (Supplemental Table 1). The percentage of EoE patches for each patient was calculated at diagnosis and remission. Model performance was assessed using the F1 score on an independent test set of 11,000 patches. Regions of interest utilized by the model for decision-making were highlighted for interpretation by pathologists. **Figure 3** shows an overview of both models.

**Figure 3.**
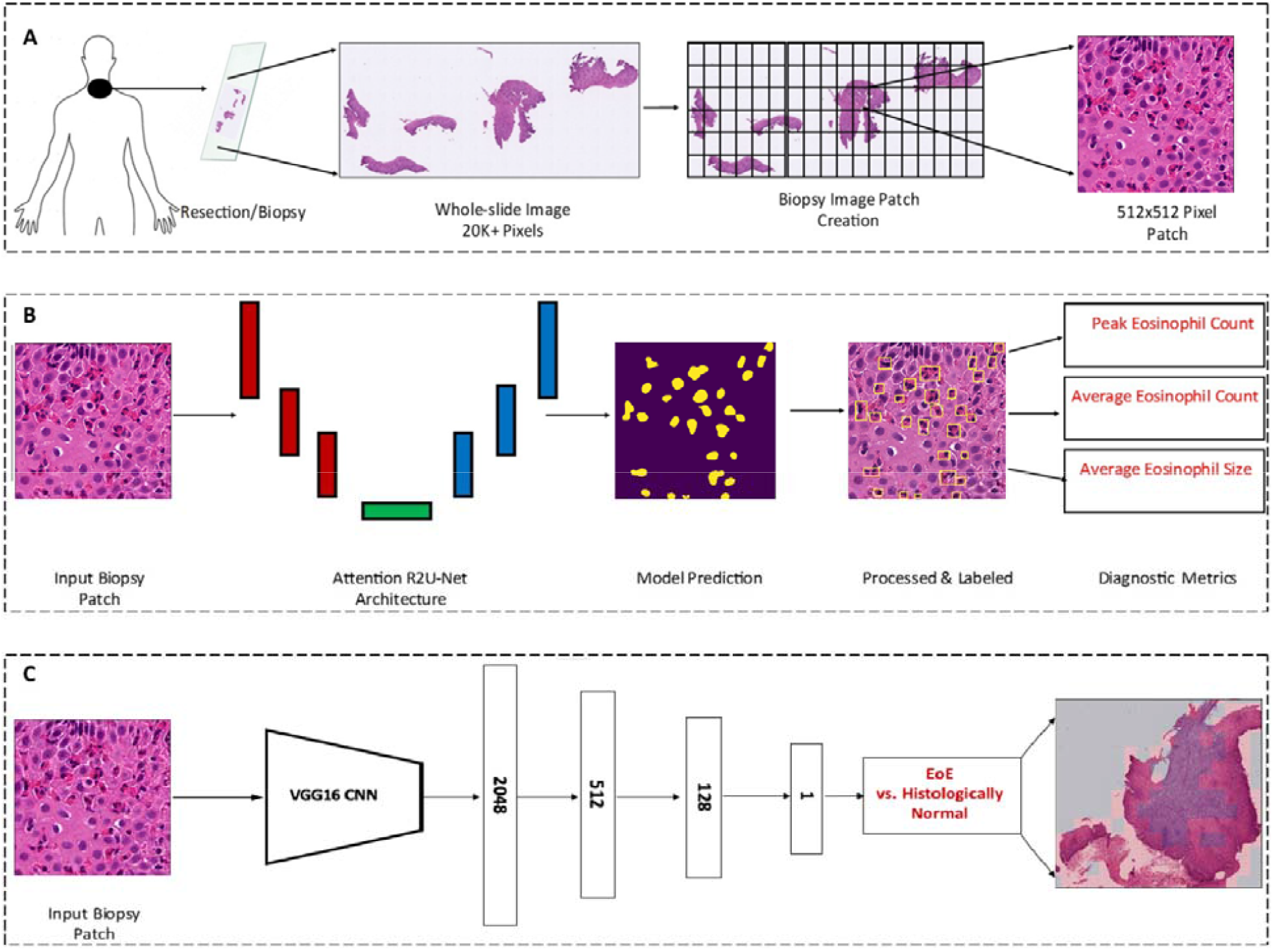
Overview of patch creation, Eo Detection Model, and EoE Classification Model. (A) Esophageal whole slide image biopsies were digitized and converted into 512×512 pixel patches for model training. (B) The Eosinophil Detection model used a U-Net segmentation architecture to identify Eos, from which HPF-level parameters were calculated such as peak Eo count, average Eo count across all HPFs in a WSI, or average Eo size across an entire WSI. (C) The EoE Classification model used a VGG-16 Convolutional Neural Network to classify patches as EoE or histologically normal. Eo = Eosinophil, EoE = Eosinophilic Esophagitis, HPF = high-powered field, WSI = whole-slide image.

### Pathology Review

Two board-certified pathologists reviewed the histology images for each patient as well as the output results for each model. All patches were reviewed and those with >10 Eo difference between true and predicted counts or Dice <0.01 (15/513 patches total, 3%) were reannotated and used to retrain the model. Classification model outputs were also reviewed and tissue patterns prevalent in areas predicted as EoE were noted.

### Evaluating associations of deep learning-derived tissue features and clinical groups

The 19 initial EoE patients were followed until they first achieved histologic remission based on biopsy reporting <15 Eos/HPF.^5^ Two patients achieved remission on pathology report but their remission biopsy WSIs could not be found. All esophageal biopsies from the initial 19 patients and the subset of 17 patients with available slides at remission were converted into patches and run through both models. The identified Eos were post-processed to extract parameters including peak Eos per HPF (PEC), average Eo count over all HPFs, average Eo size (pixels), percent of HPFs with zero Eos, percent of HPFs with <5 or 10 Eos, and percent of HPFs with ≥15, 30, 60, 90 Eos (Supplemental Figure 1). The classification model was used to calculate the percentage of patches classified as EoE as a function of a patient’s total tissue at diagnosis vs. at remission. Clinical phenotypes, treatments at time of remission biopsy, and symptoms at diagnosis and follow-up were documented. Differences in model parameters between groups were assessed using the Wilcoxon rank sum test.

## RESULTS

### EoE Cohort Clinical Characteristics

The characteristics of the total EoE cohort (n=46, 54% male, median age 33) and the subset of 19 treatment-naïve patients are shown in Table 1. Of the treatment-naïve patients, all achieved histologic remission with <15 Eos/HPF documented on pathology report over median follow-up 6 months; however, two patient’s original biopsies could not be located. At remission, all patients were on a proton-pump inhibitor (PPI), five were taking steroids and PPI, and five were following a food-elimination diet (FED) (three of the five were on FED, steroids, and PPI, while two were only on FED and PPI). All patients were symptomatic at diagnosis. Despite being in histologic remission at follow-up, nine patients were still symptomatic, six were asymptomatic, and two had no symptoms documented.

**Table 1.**
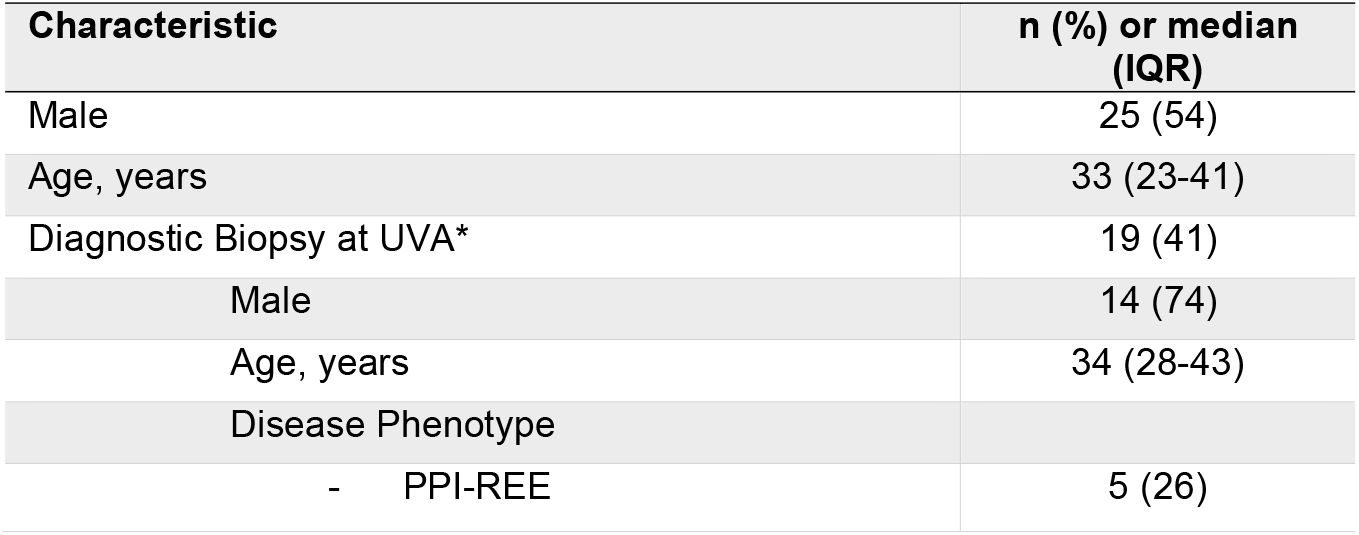
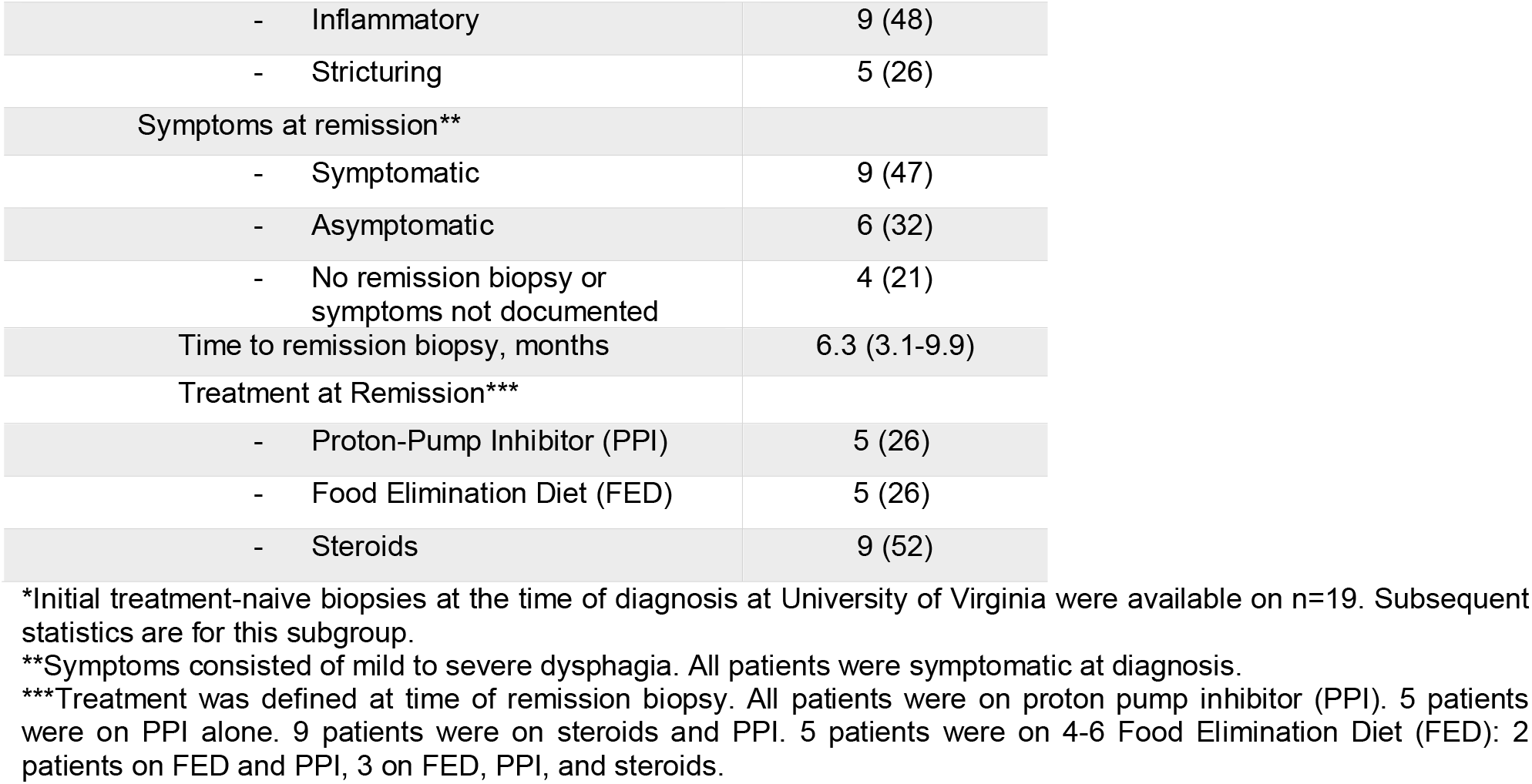
Eosinophilic Esophagitis Population Characteristics (n=46)

| Characteristic | n (%) or median (IQR) |
| --- | --- |
| Male | 25 (54) |
| Age, years | 33 (23-41) |
| Diagnostic Biopsy at UVA* | 19 (41) |
| Male | 14 (74) |
| Age, years | 34 (28-43) |
| Disease Phenotype |  |
| - PPI-REE | 5 (26) |
| - Inflammatory | 9 (48) |
| - Stricturing | 5 (26) |
| Symptoms at remission** |  |
| - Symptomatic | 9 (47) |
| - Asymptomatic | 6 (32) |
| - No remission biopsy or symptoms not documented | 4 (21) |
| Time to remission biopsy, months | 6.3 (3.1-9.9) |
| Treatment at Remission*** |  |
| - Proton-Pump Inhibitor (PPI) | 5 (26) |
| - Food Elimination Diet (FED) | 5 (26) |
| - Steroids | 9 (52) |

\*Initial treatment-naïve biopsies at the time of diagnosis at University of Virginia were available on n=19. Subsequent statistics are for this subgroup.
\*\*Symptoms consisted of mild to severe dysphagia. All patients were symptomatic at diagnosis.
\*\*\*Treatment was defined at time of remission biopsy. All patients were on proton pump inhibitor (PPI). 5 patients were on PPI alone. 9 patients were on steroids and PPI. 5 patients were on 4-6 Food Elimination Diet (FED): 2 patients on FED and PPI, 3 on FED, PPI, and steroids.

### Deep Learning Eosinophil Detection Model

The Eo detection model had a mean (SD) counting error of −0.02 (2.9) Eos/patch, or −0.3 (11.49) Eos/HPF. The Dice coefficient, or the amount of pixel overlap of predicted versus true annotations, was 72% (Supplemental Figure 2). The Pearson coefficient of predicted and true Eo counts was 0.93.

Figure 4A depicts four histograms of the total Eo distributions across all HPFs from diagnostic biopsies of four patients who were independently determined to have similar histologic severity by two gastrointestinal pathologists. The patients also had the same EoEHSS Eosinophilic Inflammation score (Grade and Stage 3) as calculated using the Eo counts generated by the model.^11^ Despite having similar histologic severity by traditional metrics, the four patients all had different PECs, which would not be captured by histologic scoring systems that stop at PEC 60 or 100 (E-23 was 100, E-92 was 150, E-127 was 200, and E-77 was 250).^8,11^ Furthermore, the distributions of Eos were vastly different between patients. For example, while patient E-77 had the highest PEC of 250, the patient also had many HPFs with 0 Eos. In comparison, E-127 had a PEC of 200 and most of their HPFs had over 50 Eos.

**Figure 4.**
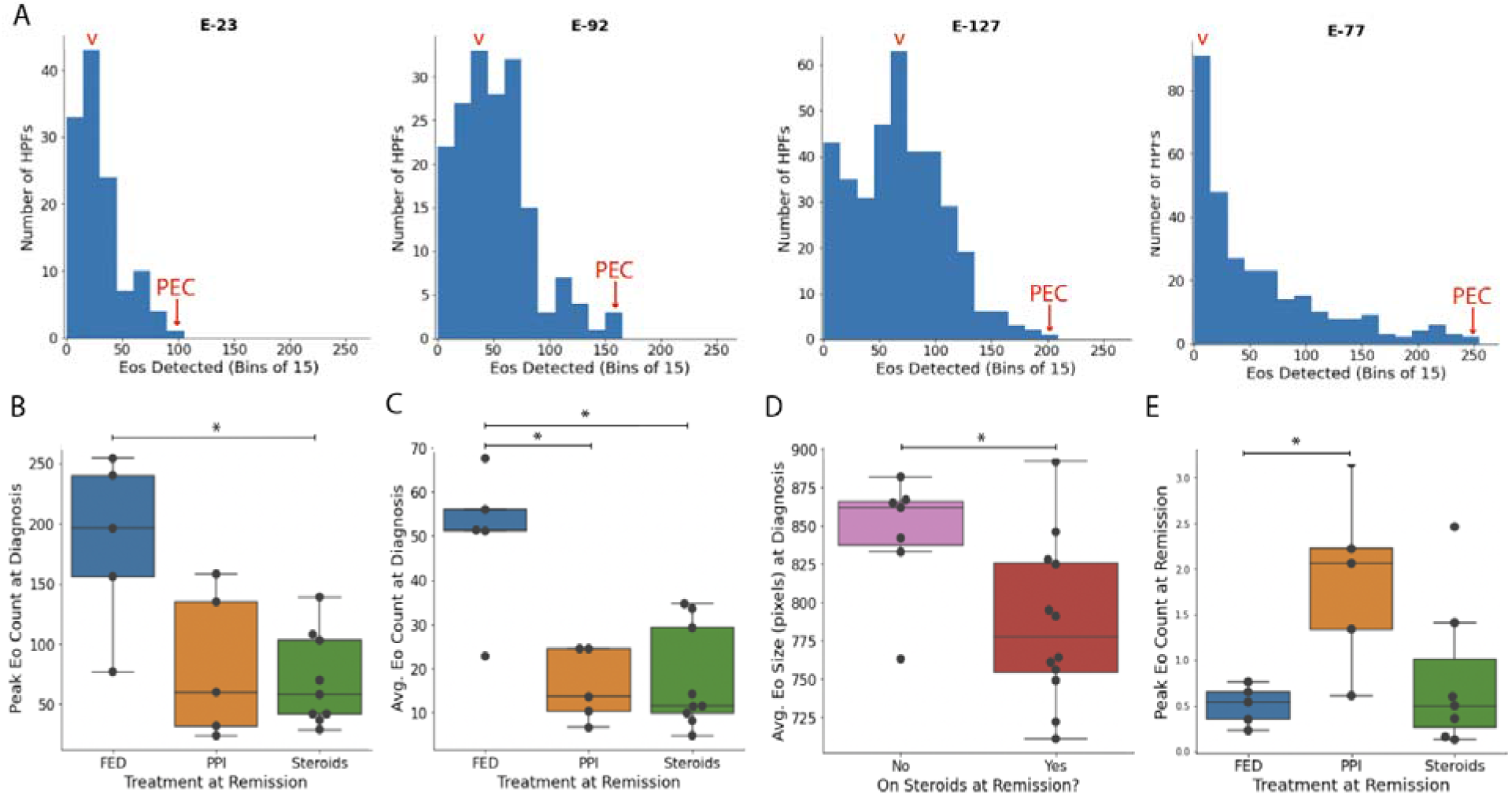
Patterns of Eo Distribution and Differences between Treatment Groups as Identified by Eo Detection Deep Learning Model. (A) Demonstration of visually different patterns in initial eosinophil (Eo) distribution among four patients (E-23, E-77, E-92, E-127) with similar histologic severity as determined independently by two board-certified gastroentestinal pathologists. Note varying modes (v) and peak Eo counts (PEC), all ≥100. (B) Patients on food elimination diet (FED) at remission had significantly higher initial PEC/high-powered field (HPF) than patients on steroids. (C) Patients on food elimination diet (FED) at remission had significantly higher initial average Eo count/HPF than patients on steroids or proton-pump inhibitor (PPI). (D) Patients who were not taking steroids at time of remission had significantly higher initial average Eo size than patients who were taking steroids. (E) Patients on FED at remission had significantly lower remission biopsy peak Eo counts than patients on PPI at remission. * indicates p<0.05

Figure 4B depicts boxplots comparing Eo parameters between groups based on the treatment they were on at remission. Patients with higher initial PECs were more likely to be on a FED than steroids at time of remission (p<0.05). Higher average Eo counts at diagnosis were also predictive of being on FED rather than steroids or PPI alone at remission (p<0.05). These trends of FED being more likely than steroids held when removing the three patients in the FED group who also received steroids (p<0.05). Patients with larger initial average eosinophil size were more likely to be on a non-steroidal therapy at time of remission (p<0.05). Upon remission, patients with lower average Eo counts were more likely to have been treated with FED than PPI alone (p<0.05). Peak eosinophil count (PEC) on initial biopsy was correlated with average eosinophil count (r=0.86) and poorly correlated with average eosinophil size (r=0.41). There were no significant associations between initial or follow-up Eo metrics and symptoms at remission.

When examining different Eo thresholds, being on FED at remission was more likely than steroids in patients with higher initial percentages of HPFs with ≥15 Eos (p <0.05), ≥30 Eos (p<0.01), ≥60 Eos (p<0.01), and ≥90 Eos (p<0.05). FED was more likely than PPI in patients with higher initial percentages of HPFs with ≥15, ≥30, and ≥60 Eos (p<0.05 for all).

### Deep Learning Eosinophilic Esophagitis Classification Model

The age and sex of histologically-normal and EoE patients are shown in Supplemental Table 1. Upon review of all WSIs from patients with <93% of their patches classified as EoE on follow-up, the pathologists identified basal zone hyperplasia and dilated intercellular spaces as features used by the models to identify EoE, which were prevalent even in tissue sections with 0 to <5 Eos (Figure 5A-B). In the test set, the EoE classification model classified 99.9% of patches from the comparison group as histologically normal and 94% of patches from EoE patients as EoE (Figure 5C). The F-1 score was 0.97.

**Figure 5.**
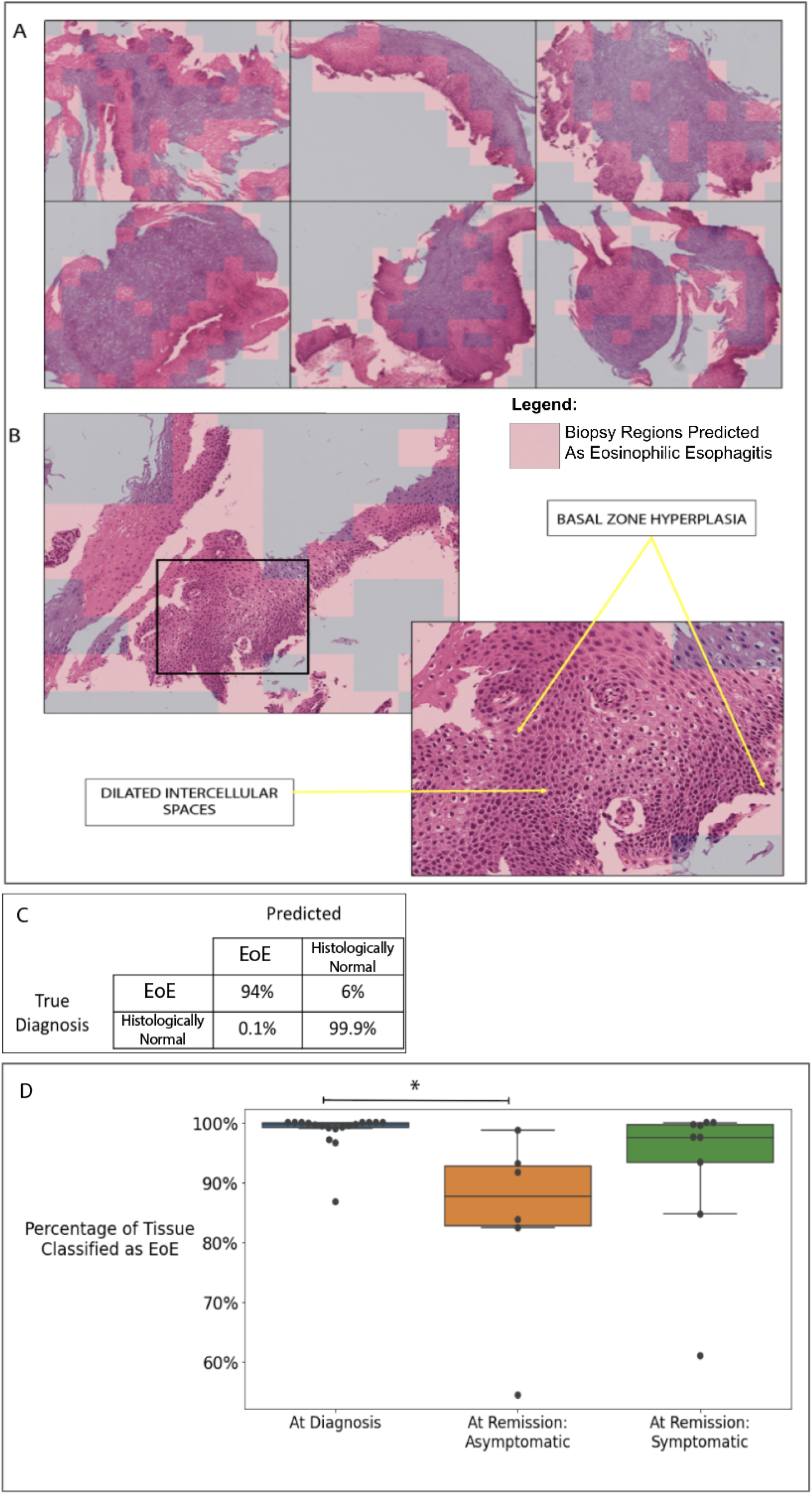
Deep Learning Eosinophilic Esophagitis Classification Model. (A) Cropped whole-slide images (WSIs) from six patients in remission with portions classified as EoE highlighted in pink. (B) Zoomed in region of one cropped WSI with arrows indicating regions with basal zone hyperplasia and dilated intercellular spaces. (C) Test Set Confusion Matrix for Classification Model. 99.9% of esophageal tissue from the comparison group was classified as histologically normal. (D) Patients who were asymptomatic at remission had significantly fewer patches classified as EoE on follow-up biopsy as compared to diagnostic biopsy. There were no significant differences between diagnostic and remission biopsies of symptomatic patients. * indicates p<0.05.

Averaged among the 19 treatment-naïve patients, the model classified 99% of diagnostic biopsy patches as EoE. Upon remission as defined by <15Eos/HPF, the model still classified 91% of patches averaged over all patients as EoE. Asymptomatic patients at remission had a significant decrease in EoE patches from diagnostic biopsies (p<0.04, Figure 5D) while symptomatic patients did not.

## DISCUSSION

We developed two deep learning approaches: first, to automate quantification of Eos throughout an entire esophageal biopsy slide – which was used to predict treatment at remission – and second, to broadly classify esophageal tissue as EoE or histologically normal – which was used to predict resolution of symptoms at follow-up. Our Eo detection model automatically detected all Eos throughout a biopsy with low mean (SD) error of −0.3 (11.5) Eos/HPF. We demonstrated considerable heterogeneity in the severity and extent of Eo infiltration among patients with the same eosinophilic inflammation severity scores.^11^ Furthermore, we were able to detect significant differences in initial and follow-up Eo counts between treatment groups, which in part was only possible because our detection model allowed for accurate quantification of PECs over 100 (and in some cases as high as 250 Eos/HPF) or average Eo counts across an entire biopsy image. Patients with the most severe Eo infiltration at diagnosis tended to be on FED over other treatments when they achieved remission. Our EoE classification model differentiated tissue from histologically normal versus EoE patients at diagnosis with 99.9% accuracy and classified 91% of tissue from patients at follow-up as EoE, despite being in remission. Upon visual review of the biopsies, the model was using features besides Eos, such as basal zone hyperplasia and dilated intercellular spaces, to identify EoE tissue. Although there were no significant associations between Eo metrics and symptoms, with a majority of patients in remission still being symptomatic, our classification model detected a significant decrease from diagnosis in EoE patches among asymptomatic patients which was not present in symptomatic patients. These findings suggest that deep learning-derived tissue features may correlate better with patient symptoms and have utility for predicting treatment sensitivity and monitoring response.

### Interpreting Eo Metrics between Treatment Groups

We found that treatment-naive patients with higher initial Eo counts were more likely to be on a FED at remission, and this is consistent with literature that suggests EoE is primarily a food-allergy mediated disease. Clayton et al. reported that EoE patients had a 45-fold increase in esophageal tissue IgG4 compared to normal patients, with abundant IgG4 antibodies to common trigger foods.^18^ Similarly, Gonsalves et al. has shown that FED significantly improves symptoms and reduces endoscopic and histopathologic features of EoE, while food reintroduction returns eosinophil counts to pre-treatment levels.^19,20^ FED has emerged as a non-pharmacologic, first-line approach for EoE management, but the specific antigens to avoid and the order of reintroduction remains an active area of investigation. Furthermore, since there are no controlled studies directly comparing diet to steroid therapy in EoE, the choice of treatment still is currently based on patient and provider preferences.^21^ Our results suggest diet therapy might be the preferred approach in people with more severe histologic eosinophilic inflammation at diagnosis. Clinical trials would be required to determine the validity of this hypothesis. The FED approach is most successful with a highly motivated patient and physician as well as available nutritionist resources.^21^

We also found that patients with larger average Eo size at diagnosis were more likely to be on non-steroidal therapy (either FED or PPI) than steroids (including patients on steroids and FED). To our knowledge, this is the first study that has investigated Eo size in EoE. In other inflammatory cell types such as neutrophils, larger cell size is related to cellular activation during an inflammatory response;^22^ however, we found Eo size was poorly correlated with other Eo metrics such as PEC or average Eo count (r=0.41). There were no significant associations with Eo size at remission and treatment groups or symptoms. Further investigation into this area may be warranted.

#### Comparisons of Tissue Features with Prior Studies

Our classification model used a similar deep learning approach to a publication earlier this year by Czyzewski et al. to classify EoE vs. histologically normal tissue;^13^ however, we have gone a step further by visualizing the features used by the model for classification and clinically correlating our findings to predict resolution of symptoms at follow-up. We noted basal cell hyperplasia and dilated intercellular spaces within regions used by the classification model to identify EoE. Basal cell hyperplasia in EoE has also been noted in several previous studies.^23,24^

Most notably, Collins et al. published two papers introducing a new EoE histological scoring system (EoEHSS)^11^ and histology remission score (EoEHRS)^8^ to grade disease severity based on eight histologic features: eosinophil density, basal zone hyperplasia, eosinophil abscesses, eosinophil surface layering, dilated intercellular spaces (DIS), surface epithelial alteration, dyskeratotic epithelial cells, and lamina propria fibrosis. They found several features in the EoEHSS, such as basal cell hyperplasia and dilated intercellular spaces, were strongly correlated with each other and significantly higher in untreated patients.^11^ They also found lower EoEHRS scores at follow-up were associated with reductions in symptoms and biomarkers for inflammatory cells such as mast cells and Eos.^8^ We have summarized the major tissue features in EoE from various papers in Supplemental Figure 3.

These scoring systems mark an important first step towards standardizing use of tissue features besides Eos for EoE patient evaluations. Our model adds several novel contributions to these scores. First, the EoEHSS evaluates PEC up to but not exceeding 60 Eos/HPF in the interest of reducing time and fatigue to produce counts, and many pathologists also stop at peak counts over 100. In contrast, our model automatically generates peak counts with no upper bound. The differences we detected in initial PECs between treatment groups could only be detected by going above 60 and in some cases over 250. Second, we detected considerable variation in the distribution of Eos among treatment-naïve patients who had the same EoEHSS Eosinophilic Inflammation score (Grade and Stage 3). Again, in the interest of saving time, these histologic scoring systems approximate the extent of inflammation throughout the biopsy while our automated model accurately quantifies Eos across all HPFs. Third, although the original authors claimed the EoEHSS scoring took <1 minute for all pathologists, our experience was that scoring took significantly longer (even generating PEC took >1 minute) and it would be difficult to evaluate a patient’s entire tissue in such a short span without potentially missing some features. In contrast, after digitizing the biopsies, the majority of our model’s analyses are performed without human input.

### Limitations

Our limitations were mostly due to limited access to data which hindered our ability to perform more powerful statistical analyses. We had diagnostic biopsies from 19 treatment-naïve EoE patients and 17 biopsies at remission. Despite a low number of patients in each treatment group, we did detect a clear trend with patients who achieved remission on FED tending to have higher Eo counts at diagnosis and lower PECs at remission. Nevertheless, these preliminary findings need to be validated in large, external datasets. Furthermore, we were only able to assess each Eo metric individually using Wilcoxon rank-sum tests to compare treatment groups. Future studies could combine multiple Eo metrics and classification results using multivariable analyses to determine if this increases the predictive power of the models. We also were not able to analyze esophageal sample location (proximal, mid, distal) due to not having sample locations for all patients. In developing the classification model, the majority of normal biopsies came from pediatric participants while the majority of EoE biopsies came from adults. It was difficult to obtain esophageal biopsies from patients with no gastrointestinal disease outside of the pediatric age range, but we have several reasons to believe this did not cause an age bias in the model. Based on the literature, there are no significant differences in the normal esophageal histology of children and adults.^25^ Furthermore, the pathophysiology and histologic findings of EoE are similar between children and adults, with the same criteria for diagnosis and remission.^1,26^ It is also possible that pediatric samples represent an idealized form of normal esophagus histology for deep learning model training, as exposure to other confounding factors such as smoking, environmental pollution, etc. is minimized. Not only were our three pediatric EoE patients in the treatment-naïve cohort classified as 99.8%, 100%, and 100% EoE at diagnosis, respectively, but also as patients got older from diagnosis to remission, the % of EoE patches went down and not up. This supports that our model is indeed classifying EoE vs. normal and not old vs. young. Lastly, our visual review of the prediction results could be considered subjective; hence, integrating these visualization techniques with approaches to quantitate cellular features may reduce human bias in model interpretation.

## CONCLUSIONS

We present two computational approaches to evaluate EoE patients using deep learning. Our Eo detection model had a low mean (SD) quantification error of −0.3 (11.5) Eos/HPF. Treatment-naïve patients with the highest peak and average Eo counts were more likely to be on FED at remission than steroids. Our EoE classification model (F1 Score 0.97) found asymptomatic patients at follow-up had a significant reduction in the percentage of tissue classified as EoE from diagnosis, while symptomatic patients had no significant differences. These results suggest that deep learning approaches may have utility in diagnosing EoE and predicting future treatment response at diagnosis and resolution of symptoms at follow-up. While our models require external validation in large and diverse cohorts, they represent a pathway to automate components of histologic analysis and potentially improve clinical assessments of EoE patients.

## Data Availability

All archival data used for this study belongs to the University of Virginia.

## Abbreviations

EoE: Eosinophilic Esophagitis
Eo: Eosinophil
UVA: University of Virginia
WSI: whole-slide image
HPF: high-powered field
FED: Food Elimination Diet
PPI: proton-pump inhibitor

## Notes

### Competing Interest Statement

The authors have declared no competing interest.

### Funding Statement

Funding Information: Research reported in this manuscript was supported by National Institute
of Diabetes and Digestive and Kidney Diseases of the National Institutes of Health under award
number K23DK117061-01A1 (Syed), University of Virginia Center for Engineering in Medicine
Grant (Syed and Brown), and University of Virginia THRIV Scholar Career Development Award
(Syed). The content is solely the responsibility of the authors and does not necessarily
represent the official views of the funding agencies.

### Author Declarations

This study was approved by the UVA Institutional Review Board (IRB #20448).

